# Governing Trust in Health AI: A Qualitative Study of Cybersecurity Professionals’ Perspectives

**DOI:** 10.64898/2026.03.01.26347389

**Authors:** Toluwani E. Adekunle, Joseph Ohaeche, Tiwaladeoluwa B. Adekunle, Deborah Adekunle, Michel Kogbe

## Abstract

**Background:** Artificial intelligence is increasingly embedded in healthcare delivery. Its legitimacy depends on institutional governance, not technical performance alone. Prior research has centered on clinicians and patients. Less attention has been given to cybersecurity professionals who sustain the digital infrastructures that support health AI. This study examines how cybersecurity professionals conceptualize AI as clinical infrastructure and how these interpretations shape understandings of trust, risk, and oversight.

**Methods:** Guided by sociotechnical systems theory and institutional trust scholarship, we conducted semi-structured in-depth interviews with twenty cybersecurity professionals working in healthcare-relevant domains. Participants were recruited through professional networks and LinkedIn outreach. Interviews were conducted between May and August 2025. They were audio-recorded and transcribed verbatim. Data were analyzed using qualitative content analysis with constant comparison. Two researchers independently coded transcripts and refined themes through iterative discussion. The study received Institutional Review Board approval.

**Results:** Participants described health AI as an augmented clinical infrastructure. They emphasized that AI extends workflow capacity but requires sustained human oversight. Healthcare data systems were characterized as fragmented and vulnerable. Breaches were treated as anticipated events. Trust in AI was described as contingent and built over time through visible accountability. Cybersecurity stewardship was framed as foundational to institutional trustworthiness.

**Conclusions:** Health AI credibility emerges through governance practices that demonstrate accountability. Cybersecurity professionals and institutional stakeholders jointly shape trust in digitally mediated healthcare systems through governance decisions that signal accountability.

## Introduction

Artificial intelligence (AI) is increasingly embedded within healthcare delivery [1],[2]. Its influence extends into clinical interpretation and operational processes [3]. Public discussion often focuses on performance and efficiency [4]. Yet credibility in healthcare depends on more than technical capability [5]. AI systems are embedded in institutional infrastructures that govern information and decision-making [6]. Sociotechnical scholarship shows that technologies gain authority through institutional embedding rather than through technical performance alone [7]. In healthcare, this embedding occurs within digital systems that remain structurally strained [8].

Cybersecurity concerns have intensified as digital dependence has grown [9]. Healthcare institutions continue to experience data breaches and ransomware attacks that expose systemic vulnerabilities [10],[11]. These incidents disrupt operations and can erode public confidence [11]. Research indicates that security failures reduce willingness to share personal health information and weaken institutional trust [12], [13]. AI systems that rely on large-scale data integration inherit underlying infrastructural weaknesses [14].

Trust in healthcare institutions has also been shaped by longer histories of inequity and harm [15]. Medical mistrust has been documented as a rational response to structural racism and unethical practices [15]. Technological expansion unfolds within this context. Algorithmic systems can reproduce inequities when trained on biased data or implemented without adequate oversight [16]. Trust in AI, therefore, reflects broader judgments about institutional responsibility [16].

Research has focused on clinicians and patients, with less attention to cybersecurity professionals who sustain AI infrastructures [9]. These actors play a critical role in shaping how risk is understood and managed [17]. Responsible innovation scholarship calls for governance to be embedded in system design rather than imposed after failure [18]. In healthcare settings, cybersecurity stewardship may signal institutional preparedness and accountability over time [18].

This study draws on sociotechnical systems theory and institutional trust scholarship. These perspectives frame technological legitimacy as an institutional achievement rather than a technical outcome [19], [20]. From this perspective, AI acquires authority through institutional governance and sustained accountability that demonstrate competence over time [21], [22].

Responsible innovation emphasizes embedding risk management into system design rather than reacting to harm [23],[24]. These perspectives position cybersecurity professionals as central to how AI-related risks are understood and addressed within healthcare organizations [25]. Fig 1.1 provides a visual representation of how cybersecurity stewardship and governance shape trust in health AI.

**Fig 1.1:**
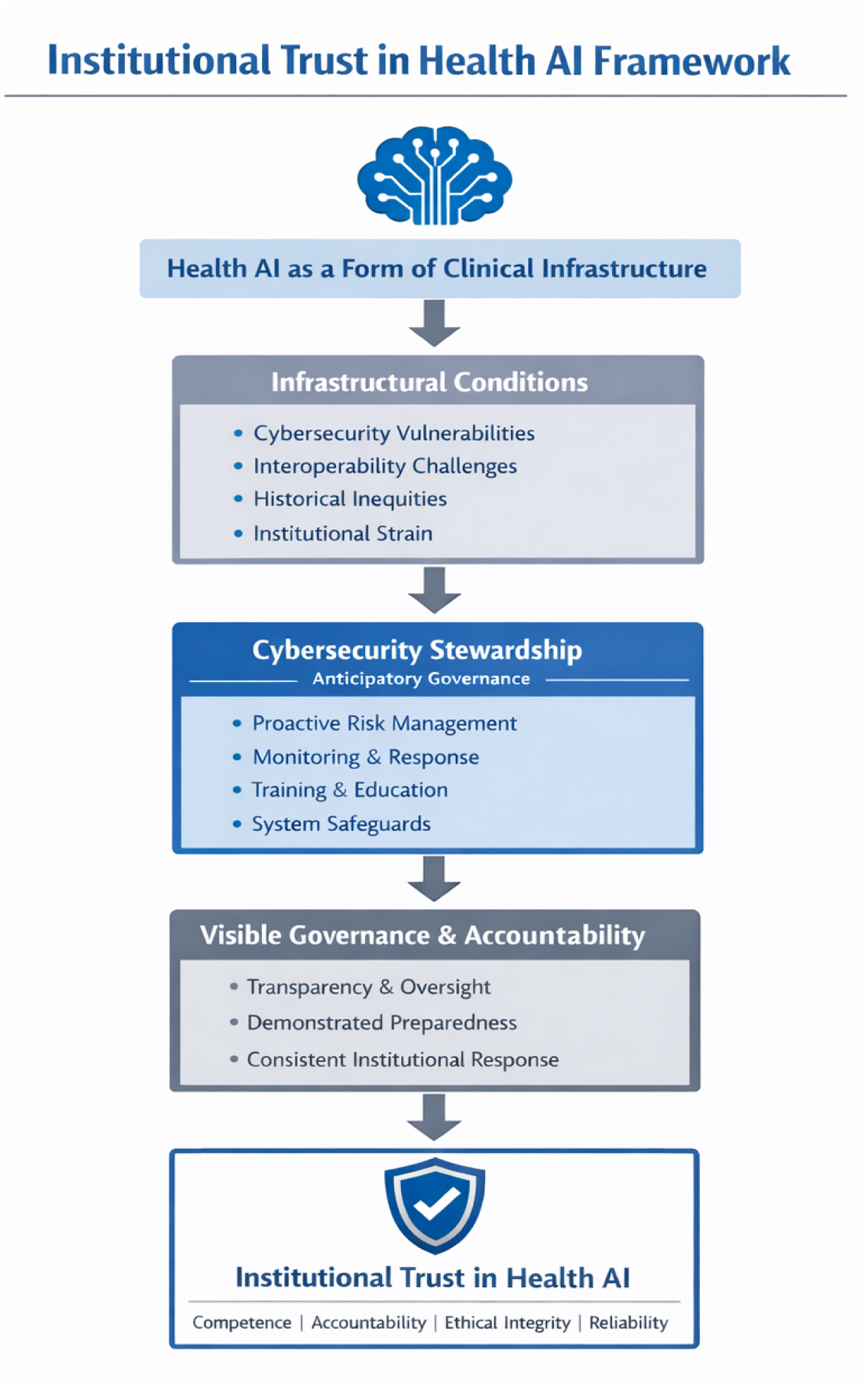
A Sociotechnical model of institutional trust in health AI

This study examines how cybersecurity professionals conceptualize artificial intelligence as clinical infrastructure and how these interpretations inform understandings of trust and oversight. By foregrounding cybersecurity perspectives, the study reframes algorithmic credibility as an institutional achievement rather than a purely technical one.

## Methods

### Study design

This qualitative study used semi-structured in-depth interviews to examine how cybersecurity professionals conceptualize artificial intelligence as clinical infrastructure and how these interpretations inform understandings of trust, risk, and oversight in healthcare systems. The study was informed by a sociotechnical framework and institutional trust theory.

### Research team and reflexivity

Interviews were conducted by the principal investigator, a public health researcher with expertise in institutional trust and health equity. The co-primary investigator is a cybersecurity professional working within a healthcare system and contributed domain expertise in digital infrastructure and risk governance. No prior relationships existed between researchers and participants. Reflexive memoing was conducted throughout data collection and analysis to examine how disciplinary perspectives informed interpretation.

### Participants and recruitment

Eligible participants were cybersecurity professionals working in domains relevant to healthcare, artificial intelligence, data systems, or digital infrastructure. Recruitment occurred through professional networks, LinkedIn outreach, and snowball sampling. This approach was selected to access specialized cybersecurity roles that are not easily reached through conventional healthcare sampling frames. Recruitment continued until thematic sufficiency was reached. Twenty participants completed interviews.

### Ethical approval

The study was approved by the Calvin University Institutional Review Board (IRB # 25-002). All participants provided written informed consent prior to participation.

### Data collection

Data were collected between May and August 2025. Interviews lasted approximately 45 to 60 minutes and were conducted via secure videoconferencing platforms. Interviews were audio recorded with permission and transcribed verbatim. Identifying information was removed during transcription. Field notes were documented following each interview.

Participants completed a brief demographic survey capturing age, race or ethnicity, education, geographic location, household income, and cybersecurity domain. Survey data were used descriptively and were not linked to quotations.

### Data analysis

Data were analyzed using qualitative content analysis to identify patterns grounded in participants’ language [26],[27]. Transcripts were read in full and coded inductively, with constant comparison used to refine conceptual categories [28]. Two researchers independently coded the data in Dedoose to support systematic organization and analytic consistency [29].

Differences in coding were resolved through discussion. Analytic memoing informed theme development [30]. Codes were abstracted into higher-order categories consistent with qualitative content analysis procedures [27]. Thematic sufficiency was reached when no substantively new categories emerged [31],[32].

### Rigor

Credibility was strengthened through interdisciplinary analytic collaboration, iterative coding, reflexive memoing, and maintenance of an audit trail [27]. Interpretations were grounded in participant quotations to support transparency and confirmability [27],[33].

## Results

Twenty participants completed the interviews. Most were ages 25 to 34 years (65%), predominantly Black or African American (80%), and held a master’s or professional degree (75%). Over half resided in urban areas (57.9%), and 45% reported household incomes of 100,000 dollars or more. Participants represented diverse cybersecurity domains, including network and infrastructure security, cloud and application security, offensive and defensive security, health informatics, data science, and artificial intelligence and machine learning.

**Table 1.1:** Demographic and professional characteristics of Cybersecurity participants (n= 20)

| **Characteristic** | **Category** | **N= 20 (%)** |
| --- | --- | --- |
| **Age** | 25-34 | 13 (65.0%) |
|  | 35-44 | 5 (25.0%) |
|  | 45-54 | 2 (10.0%) |
| **Race/Ethnicity** | Black or African American | 16 (80.0%) |
|  | Asian | 2 (10.0%) |
|  | White | 1 (5.0%) |
|  | Other | 1 (5.0%) |
| **Education** | Master's/Professional degree | 15 (75.0%) |
|  | 4 year degree | 4 (20.0%) |
|  | Doctorate | 1 (5.0%) |
| **Geographic Location** | Urban | 11 (57.9%) |
|  | Suburban | 8 (42.1%) |
| **Household Income** | \$100,000 or more | 9 (45.0%) |
| | \$50,000-\$74,999 | 5 (25.0%) |
| | \$25,000-\$49,999 | 3 (15.0%) |
| | Less than \$25,000 | 1 (5.0%) |
| | \$75,000-\$99,999 | 1 (5.0%) |
|  | Prefer not to say | 1 (5.0%) |
| **Cybersecurity domain** | Network and Infrastructure Security | 4 (22.2%) |
|  | Offensive and Defensive Security | 4 (22.2%) |
|  | Data Science and Analytics | 3 (16.7%) |
|  | Health Informatics | 3 (16.7%) |
|  | Cloud and Application Security | 2 (11.1%) |
|  | Generalist | 1 (5.6%) |
|  | Artificial Intelligence and Machine Learning | 1 (5.6%) |

Cybersecurity professionals described artificial intelligence (AI) as an increasingly embedded component of healthcare systems that both expands clinical capacity and intensifies existing vulnerabilities. Analysis revealed four interconnected patterns describing how cybersecurity professionals understand AI’s integration into clinical workflows, the fragility of surrounding data environments, the contingent nature of trust in algorithmic systems, and the organizational responsibility required to govern AI-induced clinical risk.

### Theme 1: Health AI is considered a form of augmented clinical infrastructure rather than an autonomous authority

Participants consistently described AI as extending clinical cognition and operational efficiency while rejecting its use as a replacement for human judgment. Rather than viewing AI as an independent decision maker, cybersecurity professionals framed it as infrastructure that supports clinical reasoning. One participant articulated the perceived clinical scope of AI:

> “I think AI has come to stay. I also think that AI is very beneficial in that it can analyze results, for example, X ray results, faster. It can make recommendations based on the findings. AI also helps streamline many processes. So I think in healthcare it’s good, right? Because it helps with interpreting results and medications. It also helps with finding new drugs faster as opposed to doing it manually. AI can automate several processes. And we’re also seeing the use of robots in carrying out surgeries. It’s even administering first-level care.” (Participant #001)

However, participants repeatedly emphasized that AI must operate within human-guided systems. They rejected framing AI as an autonomous authority and instead described it as a tool that augments, rather than replaces, clinical judgment. Human oversight was viewed as essential for interpreting algorithmic outputs and sustaining professional responsibility. In this framing, trust in AI depends on the continued presence of accountable human decision makers. As one participant explained:

> “Personally, I feel like there’s a good and bad side to AI, and we should not be fully reliant on it. Instead, it should be a collaboration between humans and AI. Bringing human and AI together would help in a better way because humans have the experience and the skills to analyze AI’s results. So I feel like AI is something we cannot completely rule out. Instead, we should look at how to work side by side with AI to make better decisions and achieve better results.” (Participant #002)

Participants highlighted AI’s potential to accelerate learning curves and reduce diagnostic burden. They described AI as accelerating pattern recognition and clinical expertise, particularly in image-based settings. In this framing, AI was seen as scaffolding clinical judgment rather than replacing it.

> “Using AI in the data space, I think there’s a learning curve in many of the daily processes that happen in healthcare. For example, someone in dentistry may need years of experience to pinpoint an issue in a scan. If AI can help with that, then we should explore how to make it work effectively in the healthcare space.” (Participant #003)

Several participants framed AI adoption as operationally unavoidable, particularly for efficiency and documentation. They described AI as a tool that can accelerate expertise and improve accuracy when paired with clinician review. Support was conditional, with oversight and professional judgment seen as essential. In this view, health AI functions as clinical infrastructure rather than a replacement for clinicians. This pragmatic orientation toward efficiency was captured clearly by one participant:

> “Where efficiency makes sense, you should be doing these kinds of things. It’s silly not to. As long as someone still reviews them, it’s going to be more accurate than handwritten notes. So I do think it’s a mistake not to use these tools.” (Participant #001)

Concrete examples illustrated AI’s infrastructural role in routine operations, including documentation and medication distribution. In these contexts, AI enhanced speed and reduced error, functioning as embedded operational support under institutional oversight. As one participant explained:

> “From an operational standpoint, there could be a hospital currently using AI to sort medications. They are able to deliver them from a central location massively faster and without error compared to when people were sorting the pills directly.” (Participant #005)

In these accounts, AI was embedded within backend systems that structure clinical workflow, including documentation and medication distribution. Participants emphasized precision, error reduction, and centralized coordination, framing health AI as operational infrastructure that enhances efficiency under institutional oversight.

### Theme 2: Fragile data ecologies amplify existing system vulnerabilities, deepening inequities and constraining the capacity of institutions to respond effectively

Participants consistently described healthcare data environments as fragmented and vulnerable, with AI intensifying existing structural weaknesses. Rather than portraying security risks as isolated technical failures, cybersecurity professionals emphasized system-wide fragility. One participant explained:

> “At the current stage of AI, there is extensive training and data utilization. From a cybersecurity perspective, there are concerns about whether proper procedures were followed to ensure data were not obtained through inappropriate or illegal means. There are also questions about whether the data will be stored securely, even if accessed appropriately. Overall, issues related to how data are stored, accessed, and governed, including who has access, need to be addressed.” (Participant #008)

Participants noted challenges in removing personally identifiable information across fragmented systems where data formats and storage practices vary. De-identification was described as technically complex, requiring AI to repeatedly adapt to inconsistent structures. These concerns raised broader questions about whether data governance can keep pace with heterogeneous health data infrastructures. As one participant explained:

> “Security challenges include the ability to remove personally identifiable information from patient data or results, which typically contain names and other identifiers. AI must be able to extract this information while securely storing the remaining data. However, when multiple systems store data differently, the AI has to adapt repeatedly, raising questions about whether it is consistently performing this task accurately.” (Participant #012)

Organizational preparedness for breaches was described as insufficient, with security efforts focused more on database protection than breach response. Participants viewed the lack of contingency planning as a critical gap in cybersecurity governance. As one participant explained:

> “Most systems secure their databases, but they do not prepare for a breach event. It is not enough to store and anonymize data. Given that security incidents are common, institutions should also plan for breaches so that the impact is minimized if something does happen.” (Participant #017)

Participants also raised concerns about excessive data exposure: *“There’s a tendency to give out more information to AI than necessary. And so it’s possible for someone’s information to be leaked out to the wrong personnel.”* (Participant #015)

Most systems secure their databases but do not adequately prepare for breaches. Storing and anonymizing data alone was viewed as insufficient. Participants emphasized proactive contingency planning and layered safeguards to minimize harm if exposure occurs. As one participant noted:

> “Encryption, access control, and multi factor authentication should be in place. Data storage should separate identifiable information from clinical data. That way, even if a breach occurs or information is released, only the results are visible, not the identity of the individual.” (Participant #009)

These narratives reveal AI as operating within fragile data ecologies where breaches are anticipated rather than exceptional. In this context, trust depends less on eliminating risk and more on how institutions contain harm and act responsibly within existing vulnerabilities.

### Theme 3: Contingent trust exposes the limits of algorithmic authority, as confidence in AI depends on human judgment of its relevance and reliability

Trust in AI was consistently described as partial, contingent, and mediated by human judgment. Participants resisted granting AI autonomous authority, emphasizing uncertainty related to model training, application context, and diagnostic reliability. This framing reflects an underlying recognition that algorithmic outputs are shaped by design choices and data constraints rather than objective truth. As one participant stated: “*It would depend on the AI model and the specific application, including how the model was trained.*” (Participant #005) This emphasis on model specificity illustrates how trust was not directed toward AI as a generalized category but toward particular systems operating under particular conditions.

Others explicitly limited their trust, describing it as conditional and context dependent. AI was seen as useful for routine tasks but unreliable in uncertain situations, reflecting concerns about overreliance and the limits of current training. As one participant explained::

> “It’s 50 50, very little trust. Just like doctors say not to Google your symptoms, I feel the same about AI. It has not reached a point where we can fully trust it. It works for known issues, but for unfamiliar or complex situations, it may not be helpful.” (Participant #008)

Here, participants drew parallels between AI and informal diagnostic practices, reinforcing boundaries around appropriate use and highlighting concerns about relying on AI for ambiguous or unfamiliar health issues.

Concerns about misclassification and emerging conditions were common, particularly when models may not reflect evolving clinical realities. Participants noted that healthcare knowledge changes rapidly, making reliance on static training data risky, especially if outputs are accepted without critical review. As one participant explained:

> “If the AI model is not trained properly, you might just assume that if you have these symptoms then this is the result. And with healthcare, we are learning new things every day. New diseases are everywhere. If it has not seen it before and you are relying on the results, that’s risky.” (Participant #016)

This uncertainty was compounded by awareness of technical limitations, which participants viewed as potential sources of harm rather than neutral errors.

Participants also highlighted technical failures, such as hallucinations, overdiagnosis, and underdiagnosis, as risks that warrant caution. While acknowledging AI’s current value, they emphasized that systems remain largely human-guided and should not shift toward autonomous use without careful oversight. As one participant noted:

> “The hallucinations, the generic mistakes, the overdiagnosis, the underdiagnosis… while I just said I think they’re important right now, there’s more human-guided than AI-guided per se. And so as we get closer to that, we need to be really cautious.” (Participant #007)

These concerns reinforced the necessity of continued human oversight and underscored resistance to fully delegating clinical authority to algorithmic systems. Importantly, trust was framed as relational and cumulative rather than immediate or automatic:

> “Trust is gained over time. Even if you start today, it has to be built. As people consistently receive good service and you are transparent about what you do, trust gradually grows.” (Participant #011)

These narratives suggest that trust in AI is experienced as an ongoing process. Rather than embracing AI as an authoritative replacement for human expertise, participants positioned it as a supplementary tool whose legitimacy depends on careful integration into existing clinical relationships and decision-making practices.

### Theme 4: Governing AI-induced clinical risk through cybersecurity stewardship

Participants consistently framed their professional role as extending beyond technical enforcement. As such, their roles include organizational stewardship expressed through education and institutional accountability, grounded in ethical responsibility. One participant emphasized security by design:

> “They want security built in from the beginning and integrated into discussions about what the system will do. AI systems can be vulnerable, and if security is not planned from the outset, an attacker only needs to find a single loophole to manipulate the information returned to a healthcare professional.” (Participant #012)

Cybersecurity was described as foundational infrastructure rather than a discretionary expense. Participants emphasized that failures in digital security reverberate through clinical workflows and AI generated outputs, ultimately weakening institutional trust. Investment in cybersecurity was framed as essential to maintaining system reliability and patient safety in automated care environments.

> “Cybersecurity professionals can educate stakeholders, but hospitals should not treat security as merely a cost center. It should be viewed as foundational infrastructure. Even the most advanced software is ineffective if the system crashes, as operational disruptions can directly affect what a hospital is able to do.” (Participant #017)

Participants advocated continuous monitoring and remediation, emphasizing that AI systems require ongoing oversight. Participants described it as an iterative process necessary to sustain trustworthy AI integration in clinical spaces over time.

> “They should bring in professionals to conduct penetration testing, and it should be a continuous process. Checking the system once is not enough. Six months later, you need to reassess, because systems often contain vulnerabilities that can emerge over time.” (Participant #012)

Education and awareness were positioned as critical to translating technical AI risks into organizational practice. Participants emphasize that clinicians and administrators often underestimate system vulnerabilities until breaches occur.

> “If we can streamline communication and raise awareness about how breaches occur, that would strengthen security. Many small organizations do not prioritize cybersecurity until after a breach happens. Ongoing education and training are therefore essential.” (Participant #004)

Finally, layered safeguards were emphasized as essential to minimizing harm. Participants describe encryption and access controls as mechanisms for containing damage when failures occur, rather than preventing breaches entirely.

> “Encryption, access control, and multi-factor authentication are essential to protect data. Systems should also separate identifiable information from stored data so that, if a breach occurs, the impact is minimized.” (Participant #015)

To synthesize these findings, we developed a conceptual framework that illustrates how institutional governance shapes the formation of trust in health AI (Figure 1.2).

**Fig 1.2:**
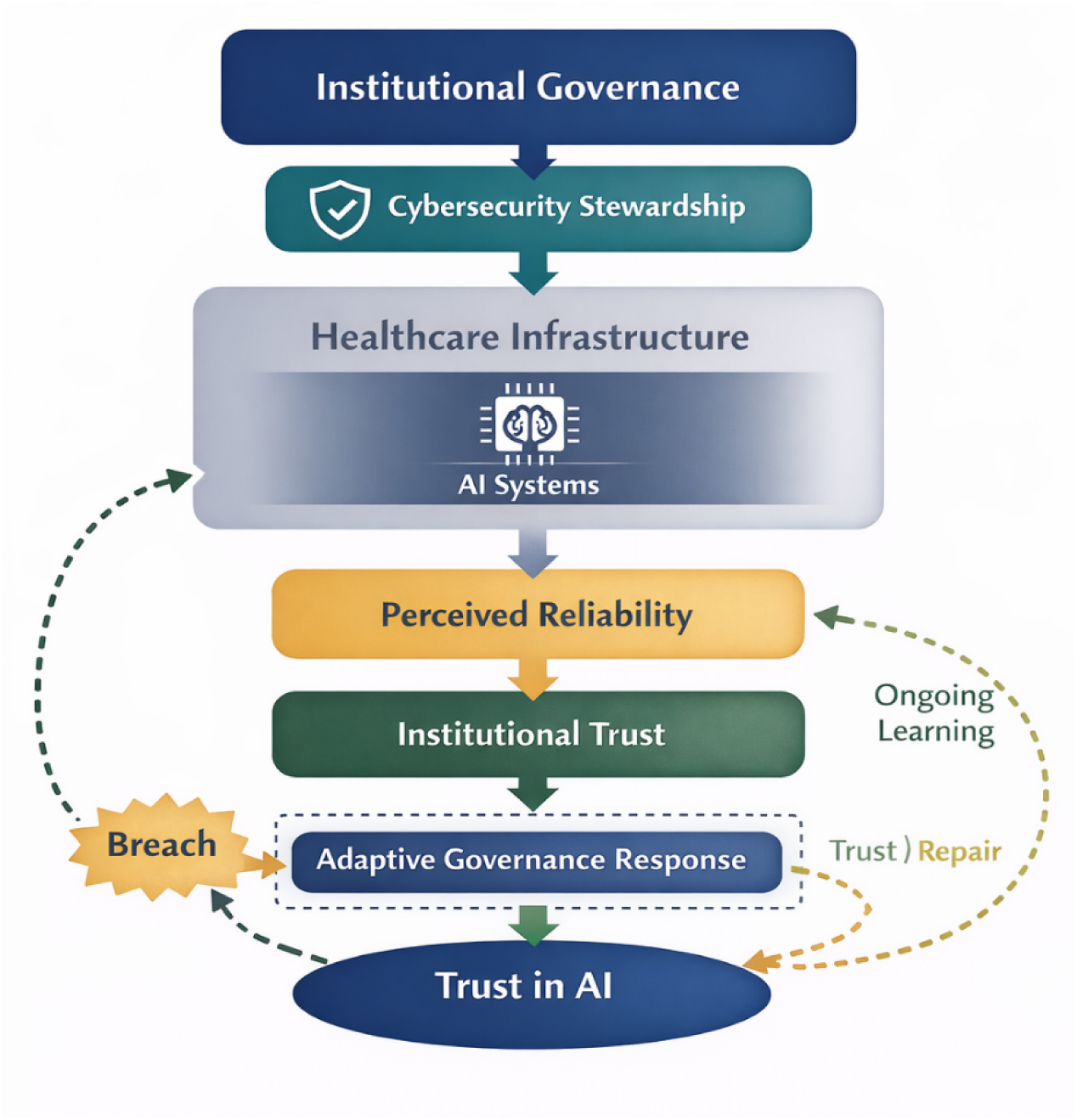
Institutional Governance and Trust Formation in Health AI: A Conceptual Model of Risk, Response, and Trust Repair

## Discussion

Cybersecurity professionals in this study did not describe artificial intelligence as an autonomous clinical authority. Instead, they spoke about it as something layered into existing systems, extending workflow capacity while remaining dependent on human interpretation. This way of thinking about AI challenges popular narratives that frame algorithmic systems as replacing professional judgment. Rather than positioning AI as an independent decision maker, participants understood it as embedded infrastructure whose legitimacy depends on continued human involvement. That perspective resonates with sociotechnical scholarship, showing that technologies acquire authority through institutional embedding rather than through technical capability alone [34], [35]. It also reflects ongoing ethical arguments in medicine that meaningful oversight cannot be outsourced to automated systems [36],[37].

The interviews also made clear that AI enters healthcare through data environments that are already strained. Participants described healthcare data systems as fragmented and unevenly governed, leaving institutions unprepared for breaches [38],[39]. These are not new concerns. Research has repeatedly shown that healthcare systems struggle with interoperability and consistent data protection practices [40],[41]. What becomes apparent in this study is how AI intensifies those vulnerabilities by relying on large-scale aggregation, retraining, and cross-system exchange. In this context, breaches were treated as expected events rather than rare anomalies, an outlook consistent with analyses of healthcare cybersecurity risk, in alignment with existing literature [18]. When institutions operate in this mode of anticipation, trust becomes tied to how well harm is contained rather than whether it is prevented [18],[42].

These infrastructural conditions matter because AI is being introduced into healthcare systems that already carry histories of inequity and distrust [43], [44], [45]. Medical mistrust is not an abstract attitudinal barrier. It has been documented as a rational response to structural racism, unethical experimentation, and persistent disparities in care [43], [44], [45]. When participants discussed data misuse, opaque training processes, or insufficient safeguards, their concerns intersected with these broader histories [46],[25]. Emerging evidence suggests that algorithmic tools can reproduce inequities when the underlying data reflect structural bias [47],[48]. In such settings, AI systems inherit the institutional reputation of the organizations deploying them.

Trust in health AI was described by participants as situation-specific and built over time. Confidence depended on understanding how models were trained, what kinds of data informed them, and whether outputs remained subject to clinical review. This understanding aligns with longstanding theoretical accounts of trust as relational and iterative rather than fixed [49],[50]. It also parallels research showing that institutional trust develops through repeated experiences of transparency and accountability [51],[52]. In other words, trust in AI cannot be separated from trust in the systems that surround it [53],[54]. If oversight appears thin or reactive, skepticism follows. If governance is visible and consistent, confidence grows gradually [55].

Participants also spoke about their own roles in ways that extended beyond technical maintenance. They described embedding security from the outset, advocating for continuous testing, and educating colleagues about digital risk. These processes were described as collaborative, involving cybersecurity expertise alongside institutional stakeholders such as clinical leadership, administrators, and legal teams who ultimately determine risk tolerance and operational accountability. These practices resemble what responsible innovation scholars describe as anticipatory governance, in which risk management is integrated into design rather than appended after harm occurs [56],[57]. The implication is that cybersecurity work is not peripheral to patient trust. It is part of how institutions signal responsibility. Evidence from breach research shows that data security failures can weaken patient confidence and engagement [58],[59]. From this vantage point, cybersecurity stewardship becomes intertwined with institutional trustworthiness.

### Implications

This study highlights cybersecurity professionals and institutional stakeholders as central to how health AI is governed in practice. Their accounts suggest that the credibility of algorithmic systems depends not only on technical performance but on the institutional environments in which they operate and on organizations’ demonstrated capacity to manage risk over time. Governance was described as ongoing institutional work rather than a fixed set of safeguards, making trust in AI closely tied to perceptions of organizational preparedness and accountability.

The findings also indicate that AI adoption cannot be separated from the reputational standing of the institutions deploying it. When AI is introduced into data environments already perceived as fragile, confidence in technological systems becomes intertwined with confidence in organizational competence. In such contexts, governance practices shape how technological expansion is interpreted socially, particularly where prior harms have complicated institutional relationships with communities.

Participants further described cybersecurity work as embedded within system development and everyday organizational activity rather than applied only after failure. This positioning aligns with responsible innovation perspectives that treat risk management as integral to design and suggests that cybersecurity stewardship contributes directly to institutional trustworthiness.

### Limitations

This study examines how cybersecurity professionals understand the integration of AI into healthcare infrastructure. Their accounts show how governance is interpreted by those responsible for system integrity. They do not capture how these practices are perceived by patients or clinicians. Institutional preparedness may be experienced differently in clinical settings, especially among communities whose trust has been shaped by prior harm. Future research should examine whether these governance practices are visible or meaningful beyond technical roles.

The findings reflect professional interpretation rather than direct observation of institutional practice. Participants described experiences shaped by specific organizational and technological contexts. Healthcare systems differ substantially across settings, so these findings should be understood as interpretive rather than representative of all health AI governance environments.

The study also captures perspectives during a period of rapid technological and security change. Governance practices may evolve as institutions respond to new risks and regulatory expectations. Longitudinal research is needed to understand how professional views of responsibility and oversight shift as health AI becomes more embedded in clinical infrastructure.

Finally, the study examines how professionals conceptually link governance and trust. It does not measure whether governance practices change patient behavior, engagement, or institutional reputation. Future empirical work should examine how cybersecurity stewardship influences trust related outcomes in healthcare over time.

## Conclusion

Health AI is often evaluated in terms of performance or efficiency. This study shows that its credibility depends on institutional context and governance. Cybersecurity professionals described AI as embedded within infrastructures that require sustained oversight and accountability. Trust develops when institutions demonstrate their capacity to manage risk and respond to vulnerability over time. In healthcare environments shaped by prior harm and uneven confidence, strengthening cybersecurity governance is therefore more than technical protection. It is part of how institutions establish and maintain trust as digital systems become integral to clinical care.

## Data Availability

The data underlying the findings of this study consist of qualitative interview transcripts with cybersecurity professionals. Due to the potential for identification of participants and the ethical restrictions imposed by the Institutional Review Board (Calvin University IRB #25-002), the full transcripts cannot be made publicly available. Deidentified excerpts supporting the findings are included within the manuscript. Qualified researchers may request access to additional deidentified data from the corresponding author, subject to institutional data use agreements and approval by the Institutional Review Board.

